## Supplementary material for "Impact of prior SARS-CoV-2 infection on perioperative cardiac, pulmonary and neurocognitive complications in elderly patients: study protocol for an observative cohort study": SPIRIT-Fig1

**Figure 1** : Timetable

| | STUDY PERIOD | | | | | | |
|---|---|---|---|---|---|---|---|
| | Baseline | Day of surgery | Postoperative Day | | | | |
| **TIMEPOINT** | Pre-surgery | 0 | *1* | *2* | *3* | *4* | *5* |
| **ENROLMENT:** | | | | | | | |
| **In- and exclusion criteria, consent** | x | | | | | | |
| **Demographic data** | x | | | | | | |
| **Physical status (CCI, iADL)** | x | | | | | | |
| **Main diagnosis, Type of surgery** | x | | | | | | |
| **Medication** | x | | | | | | |
| **Frailty (CFS and Fried criteria)** | x | | | | | | |
| **MOCA** | x | | | | | | |
| **COVID-Status** | x | | | | | | |
| **INTERVENTIONS:** | | | | | | | |
| **Surgery procedure** | | x | | | | | |
| **Laboratory parameters** | x | x | | | | | |
| **Anesthesia data** | | x | | | | | |
| **ASSESSMENTS:** | | | | | | | |
| **Cardial complications** | | x | x | x | x | x | x |
| **Pulmonal complications** | | x | x | x | x | x | x |
| **Delirium (4AT und 3DCAM)** | | x | x | x | x | x | x |
| **Other complications (PONV)** | | x | x | x | x | x | x |
| **Patients' questionnaire (pain)** | | x | x | x | x | x | x |
| **Extraction of clinical data** | x | x | x | x | x | x | x |
| **Use of interim reports** | | x | x | x | x | x | x |
| **X-Ray, MRI, CT-Reports** | | x | x | x | x | x | x |

* *CCI* Charlson Comorbidity Index, *iADL* instrumental activities of daily living, *CFS* clinical frailty scale, SF-36 short-form assessment of *MOCA* Montreal Cognitive Assessment, *4AT* 4A´s Test for delirium and cognitive decline, *3DCAM* 3-dimensional Confusion Assessment Method, *PONV* Postoperative Nausea und Vomiting
